# A Two-Sample Robust Bayesian Mendelian Randomization Method Accounting for Linkage Disequilibrium and Idiosyncratic Pleiotropy with Applications to the COVID-19 Outcome

**DOI:** 10.1101/2021.03.02.21252801

**Authors:** Anqi Wang, Zhonghua Liu

## Abstract

Mendelian randomization (MR) is a statistical method exploiting genetic variants as instrumental variables to estimate the causal effect of modifiable risk factors on an outcome of interest. Despite wide uses of various popular two-sample MR methods based on genome-wide association study summary level data, however, those methods could suffer from potential power loss or/and biased inference when the chosen genetic variants are in linkage disequilibrium (LD), and also have relatively large direct effects on the outcome whose distribution might be heavy-tailed which is commonly referred to as the idiosyncratic pleiotropy phenomenon. To resolve those two issues, we propose a novel Robust Bayesian Mendelian Randomization (RBMR) model that uses the more robust multivariate generalized *t*-distribution (Arellano-Valle and Bolfarine, 1995) to model such direct effects in a probabilistic model framework which can also incorporate the LD structure explicitly. The generalized *t*-distribution can be represented as a Gaussian scaled mixture so that our model parameters can be estimated by the EM-type algorithms. We compute the standard errors by calibrating the evidence lower bound using the likelihood ratio test. Through extensive simulation studies, we show that our RBMR has robust performance compared to other competing methods. We also apply our RBMR method to two benchmark data sets and find that RBMR has smaller bias and standard errors. Using our proposed RBMR method, we find that coronary artery disease is associated with increased risk of critically ill coronavirus disease 2019 (COVID-19). We also develop a user-friendly R package *RBMR* (https://github.com/AnqiWang2021/RBMR) for public use.

## 1. Introduction

Mendelian randomization (MR) is a useful statistical method that leverages genetic variants as instrumental variables (IVs) for assessing the causal effect of a modifiable risk factor on a health outcome of interest even in the presence of unmeasured confounding factors (Ebrahim and Smith, 2008; Lawlor et al., 2008; Evans and Davey Smith, 2015). Because of the inborn nature of genetic variants, the associations between genetic variants and phenotypes after adjusting for possible population stratification will not be confounded by the environmental factors, socio-economic status and life styles after birth. Genome-wide association studies (GWAS) have identified tens of thousands of common genetic variants associated with thousands of complex traits and diseases (MacArthur et al., 2017). Those GWAS summary level data contain rich information about genotype-phenotype associations (https://www.ebi.ac.uk/gwas/), and thus provide us valuable resources for MR studies. Therefore, we have seen a boost of two-sample MR method developments and applications based on GWAS summary statistics recently due to the increasing availability of candidate genetic variant IVs for thousands of phenotypes. (Burgess et al., 2013; Bowden et al., 2015; Pickrell et al., 2016). In particular, a genetic variant serving as a valid IV must satisfy the following three core assumptions (Martens et al., 2006; Lawlor et al., 2008):

1. **Relevance:** The genetic variant must be associated (not necessarily causally) with the exposure;
2. **Effective Random Assignment:** The genetic variant must be independent of any (measured or unmeasured) confounders of the exposure-outcome relationship;
3. **Exclusion Restriction:** The genetic variant must affect the outcome only through the exposure, that is, the genetic variant must have no direct effect on the outcome not mediated by the exposure.

When these three core IV assumptions hold, the inverse variance weighted (IVW) (Ehret et al., 2011) method can be simply used to obtain unbiased causal effect estimate of the exposure on the outcome. However, among those three core assumptions, only the IV relevance assumption can be empirically tested, for example, by checking the empirical association strength between the candidate IV and the exposure using the GWAS catalog (https://www.ebi.ac.uk/gwas/). The association between the IV and the exposure must be strong enough (the IV explains a large amount of the variation of the exposure variable) to ensure unbiased causal effect estimate. The problem of weak IVs has been studied previously in the econometric literature (Bound et al., 1995; Hansen et al., 2008). In MR settings, the method that uses genetic score by combining multiple weak IVs together to increase the IV-exposure association strength to reduce weak IV bias has also been proposed (Evans et al., 2013). Unfortunately, the other two IV core assumptions cannot be empirically tested and might be violated in practice. Violation of the exclusion restriction assumption can occur when the genetic variant indeed has a non-null direct effect on the outcome not mediated by the exposure, referred to as systematic pleiotropy (Solovieff et al., 2013; Verbanck et al., 2018; Zhao et al., 2020b). However, very often, genetic variants might have relatively large direct effects whose distribution exhibit heavy-tailed pattern, a phenomenon referred to as the idiosyncratic pleiotropy in this paper.

To address those possible violations of the IV core assumptions and potential risk, many efforts have been made recently. The MR-Egger regression method introduced an intercept term to capture the presence of unbalanced systematic pleiotropy under the Instrument Strength Independent of Direct Effect (InSIDE) assumption (Bowden et al., 2015). However, MR-Egger would be biased when there exists idiosyncratic pleiotropy. Zhu et al. (2018) proposed the GSMR method that removes suspected genetic variants with relatively large direct effects and also takes the LD structure into account by using the generalized least squares approach. However, removal of a large number of relatively large direct effects might lead to efficiency loss. Zhao et al. (2020b) proposed MR-RAPS to improve statistical power for causal inference and limit the influence of relatively large direct effects by using the adjusted profile likelihood and robust loss functions assuming that those SNP IVs are independent. However, this independent IV assumption might not hold in practice because SNPs within proximity tend to be correlated. Cheng et al. (2020) proposed a two-sample MR method named MR-LDP that built a Bayesian probabilistic model accounting for systematic pleiotropy and LD structures among SNP IVs. One drawback of the MR-LDP method is that it cannot handle relatively large direct effects well.

To overcome the limitations of those aforementioned methods, we propose a more robust method named ‘Robust Bayesian Mendelian Randomization (RBMR)’ accounting for LD, systematic and idiosyncratic pleiotropy simultaneously in a unified framework. Specifically, to account for LD, we first estimate the LD correlation matrix of SNP IVs and then explicitly include it in the model likelihood. To account for idiosyncratic pleiotropy, we propose to model the direct effects using the more robust multivariate generalized *t*-distribution (Arellano-Valle and Bolfarine, 1995; Frahm, 2004) which will be shown to have improved performance than using the Gaussian distribution when the idiosyncratic pleiotropy is present. Moreover, this more robust distribution can be represented as a Gaussian scaled mixture to facilitate model parameter estimation using the parameter expanded variational Bayesian expectation maximization algorithm (PX-VBEM) (Yang et al., 2020) which combines the VB-EM (Beal et al., 2003) and the PX-EM (Liu et al., 1998) together. We further calculate the standard error by calibrating the evidence lower bound (ELBO) according to a nice property of the likelihood ratio test (LRT). Both extensive simulation studies in Section 3 and analysis of two real benchmark data sets in Section 4 show that our proposed RBMR method outperforms competitors. We also find that coronary artery disease (CAD) is associated with increased risk of critically ill COVID-19 outcome.

## 2. Methods

### 2.1 The Linear Structural Model

Suppose that we have *J* possibly correlated genetic variants (for example, single-nucleotide polymorphisms, or SNPs) *G*_*j*_, *j* = 1, 2, …, *J*, the exposure variable *X*, the outcome variable *Y* of interest and unknown confounding factors *U*. Let *δ*_*X*_ and *δ*_*Y*_ denote the effects of confounders *U* on exposure *X* and outcome *Y* respectively. The coefficients *γ*_*j*_ (*j* = 1, 2, …, *J*) denote the SNP-exposure true effects. Suppose that all the IVs are valid, then the exposure can be represented as a linear structural function of the SNPs, confounders and an independent random noise term *e*_*X*_. The outcome can be represented as a linear structural function of the exposure, confounders and the independent random noise term *e*_*Y*_. The true effect size of the exposure on the outcome is denoted as *β*_0_. Then, we have the following linear structural equation models (Bowden et al., 2015):

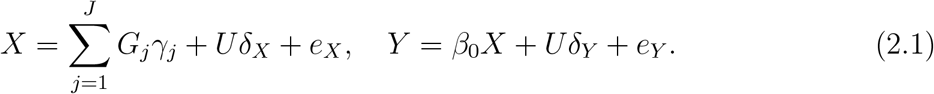

Let Γ_*j*_ (*j* = 1, 2, …, *J*) be the true effects of SNPs on the outcome. With valid IVs, we have

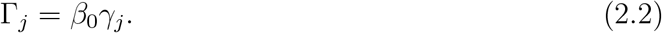

To accommodate possible violations of the exclusion restriction assumption, we now consider the following modified linear structural functions (Bowden et al., 2015):

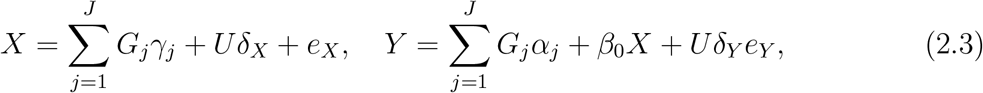

where the coefficients *α*_*j*_ (*j* = 1, 2, …, *J*) represent the direct effects of the SNPs on the outcome. Then we have

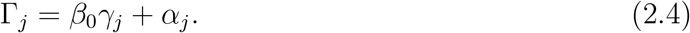

So far, many existing MR methods assign the Gaussian distribution on each direct effect *α*_*j*_, that is 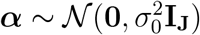 (Zhao et al., 2020b; Cheng et al., 2020; Zhao et al., 2020a), where ***α*** = [*α*_1_, …, *α*_*J*_]^T^ is a *J*-dimensional vector of direct effects. However, real genetic data might contain some relatively large direct effects whose distribution can be heavy-tailed, and thus the Gaussian distribution might not be a good fit. Therefore, we propose to assign the multivariate generalized *t*-distribution on ***α*** (Arellano-Valle and Bolfarine, 1995; Kotz and Nadarajah, 2004), which is a robust alternative to the Gaussian distribution (Frahm, 2004).

### 2.2 The Robust Bayesian MR Model

Let 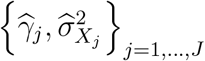 and 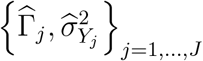 be the GWAS summary statistics for the exposure and the outcome respectively, where 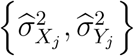 are the corresponding estimated standard errors. Many existing MR methods assume that IVs are independent from each other (Ehret et al., 2011; Bowden et al., 2015; Zhao et al., 2020b), and the uncorrelated SNPs can be chosen by using a tool called LD clumping (Hemani et al., 2016; Purcell et al., 2007), which might remove many SNP IVs and thus cause efficiency loss. To include more SNP IVs even if they are in LD, we need to account for the LD structure explicitly. To achieve this goal, we use a reference panel sample to assist with reconstructing LD matrix, such as the 1000 Genome Project Phase 1 (*N* =379) (Consortium et al., 2012). We first apply the LDetect method to partition the whole genome into *Q* blocks (Berisa and Pickrell, 2016) and then estimate the LD matrix **Θ** using the estimator 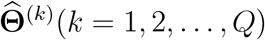 first proposed by Rothman (2012). Then, the distributions of 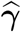 and 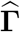 are given by

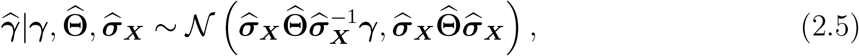

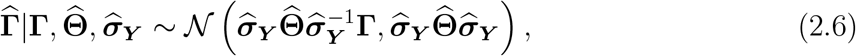

where 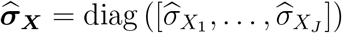 and 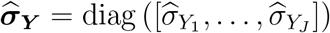 are both diagonal matrices (Zhu and Stephens, 2017).

To account for the presence of idiosyncratic pleiotropy, we propose to model the direct effects ***α*** using the more robust multivariate generalized *t*-distribution (Arellano-Valle and Bolfarine, 1995; Kotz and Nadarajah, 2004; Ala-Luhtala and Piché, 2016) whose density function is given by

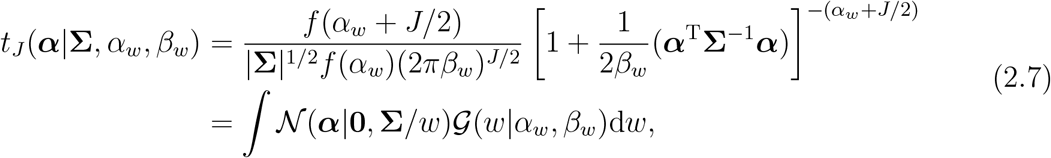

where 𝒩 (***α***|**0, Σ***/w*) denotes the *J*-dimensional Gaussian distribution with mean **0** and covariance 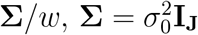 is a *J* × *J* diagonal matrix, and *𝒢* (*w*|*α*_*w*_, *β*_*w*_) is the Gamma distribution of a univariate positive variable *w* referred to as a weight variable

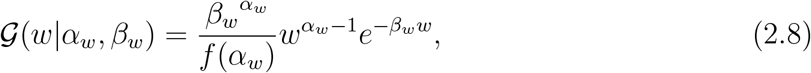

where *f* denotes the Gamma function. When *α*_*w*_ = *β*_*w*_ = *ν/*2 in equation (2.8), the distribution in equation (2.7) reduces to a multivariate *t*-distribution, where *ν* is the degree of freedom. Gaussian scaled mixture representation enables the use of EM-type algorithms for statistical inference, such as the PX-VBEM (Yang et al., 2020) described in Section 2.3. Then we denote the distribution of the latent variable ***γ*** as

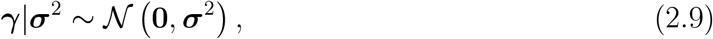

where ***σ***^2^ = *σ*^2^**I**_**J**_ is a *J* × *J* diagonal matrix. By assuming that ***γ, α*** and *w* are latent variables, the complete data likelihood can be written as

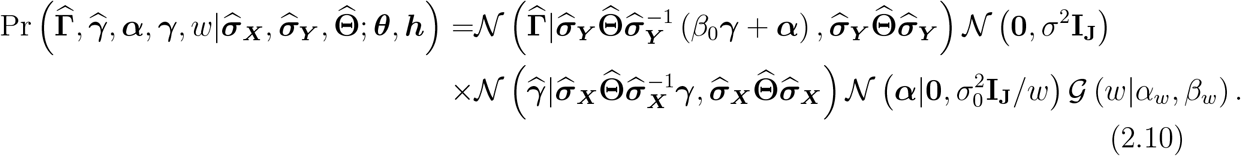

### 2.3 Estimation and Inference

The standard expectation-maximization (EM) algorithm (Dempster et al., 1977) is a popular choice for finding the maximum likelihood estimate in the presence of missing (latent) variables. However, one difficulty for implementing the EM algorithm is to calculate the marginal likelihood function which might involve difficult integration with respect to the distributions of the latent variables. In addition, the original EM algorithm might be slow (Liu et al., 1998). To address these numerical issues, we utilize a parameter expanded variational Bayesian expectation-maximization algorithm, namely, PX-VBEM (Yang et al., 2020), by replacing the EM algorithm in VB-EM (Beal et al., 2003) with PX-EM algorithm (Liu et al., 1998) to accelerate the speed of convergence. To start with, for the purpose of applying the PX-EM algorithm, the distribution of 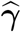 in equation (2.5) can be rewritten as follows:

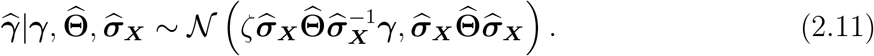

We also rewrite the complete data likelihood in equation (2.10) as:

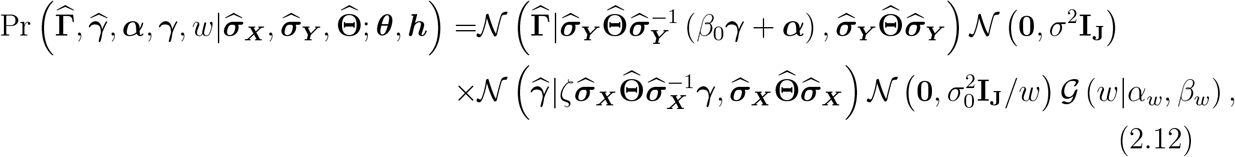

where the expanded model parameters for RBMR are 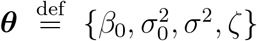. Let *q*(***γ, α***, *w*) be a variational posterior distribution. The logarithm of the marginal likelihood can be decomposed into two parts,

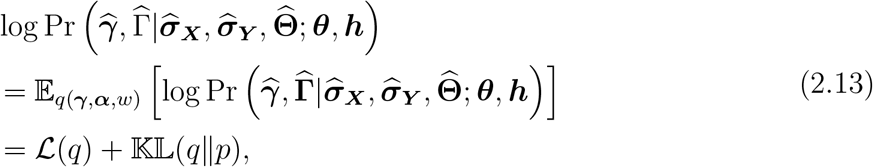

Where

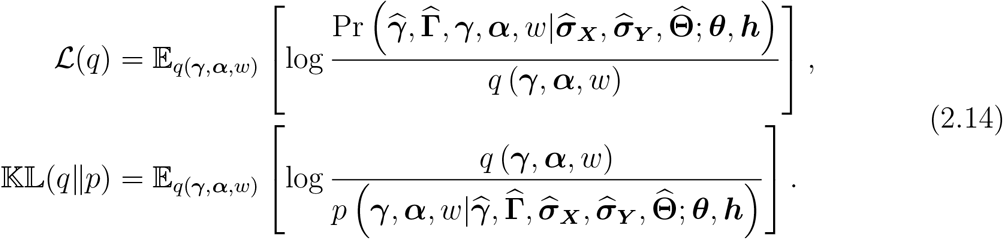

Given that the *ℒ* (*q*) is an evidence lower bound (ELBO) of the marginal log-likelihood, the non-negative Kullback-Leibler (KL) divergence 𝕂 𝕃 (*q*‖/*p*) is equal to zero if and only if the variational posterior distribution is equal to the true posterior distribution. Minimizing the KL divergence is equivalent to maximizing ELBO. Before calculating the maximization of ELBO, due to the fact that latent variables are independent of each other, the decomposition form of the posterior distribution *q*(***γ, α***, *w*) is obtained using the mean field assumption (Blei et al., 2017),

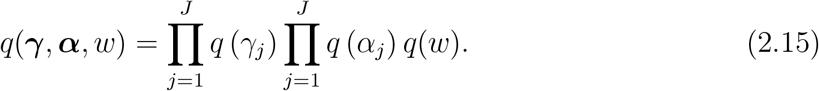

In the PX-VB-E step, the optimal variational posterior distributions for ***γ, α*** and *w* can be written as:

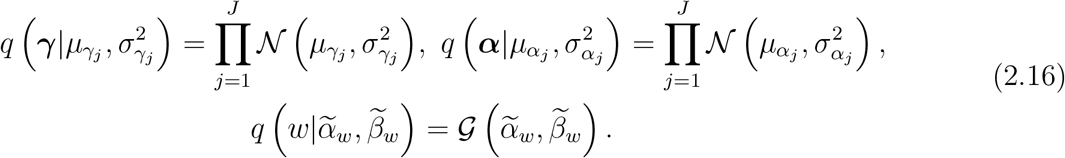

The updating equations for the parameters are given by

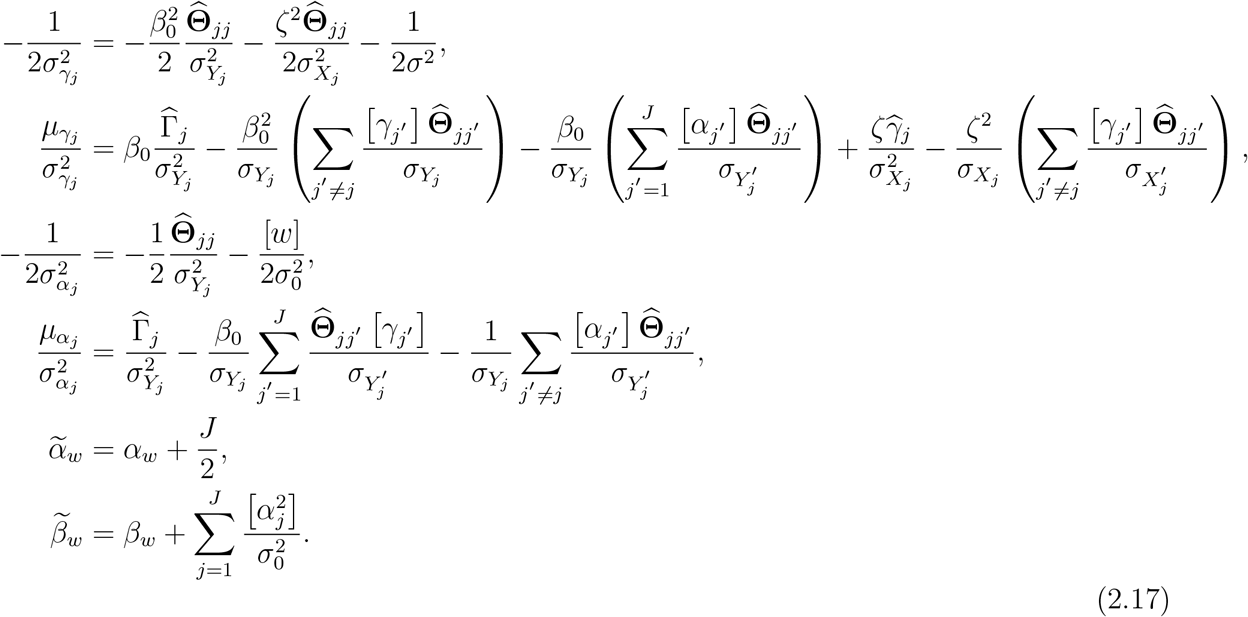

where 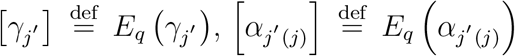 and 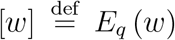.

In the PX-VB-M step, by setting the derivate of the ELBO to be zero, the model parameters ***θ*** can be obtained as:

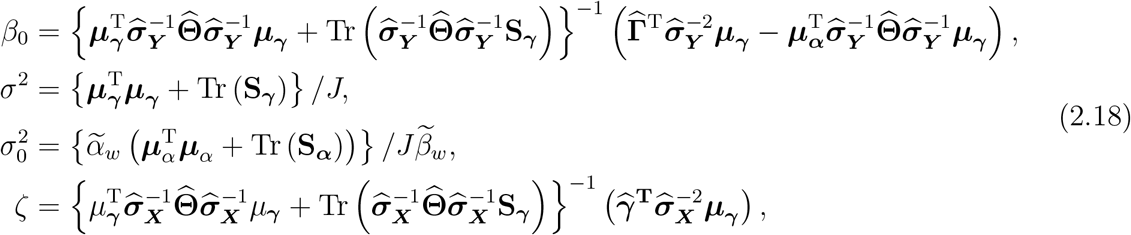

Where 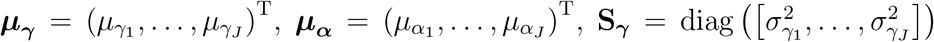 and 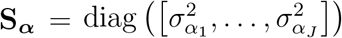. Finally, we use the updated model parameters ***θ*** to construct the evidence lower bound to check the convergence. Since we adopt PX-EM algorithm, the reduction step should be used to process the obtained parameters. More technical details can be found in the Supplementary Materials.

After obtaining an estimate of the causal effect, we further calculate the standard error according to the property of likelihood ratio test (LRT) statistics which asymptotically follows the 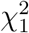 under the null hypothesis (Van der Vaart, 2000). We first formulate the statistical tests to examine the association between the risk factor and the outcome.

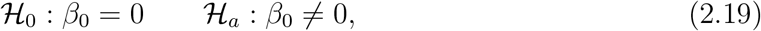

the likelihood ratio test (LRT) statistics for the causal effect is given by:

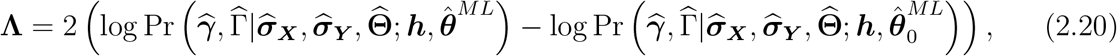

Where 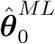 and 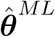 are collections of parameter estimates obtained by maximizing the marginal likelihood under the null hypothesis *ℋ*_0_ and under the alternative hypothesis *ℋ*_*a*_. We utilize PX-VBEM algorithm to maximize the ELBO to get the 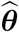 and 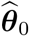 instead of maximizing the marginal likelihood to overcome the computational intractability. Although PX-VBEM produces accurate posterior mean estimates (Blei et al., 2017; Dai et al., 2017; Yang et al., 2018), it would underestimate the marginal variance because we use the estimated posterior distribution from the ELBO to approximate the marginal likelihood in equation (2.20) (Wang and Titterington, 2005). Thus, we calibrate ELBO by plugging our estimates (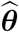 and 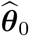) from PX-VBEM into the equation (2.20) to construct the test statistics (Yang et al., 2020):

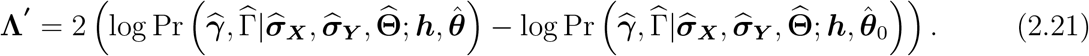

Then, we can get the well-calibrated standard error as 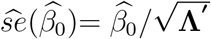.

## 3. Simulation Studies

Although our proposed method is based on summary level data, we still simulate the individual-level data to better mimic real genetic data sets. Specifically, the data sets are generated according to the following models:

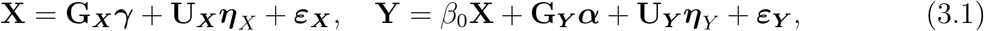

Where 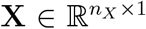 is the exposure vector, 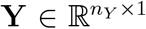 is the outcome vector, 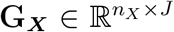 and 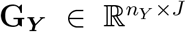 are the genotype datasets for the exposure **X** and the outcome **Y**, 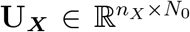 and 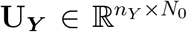 are matrices for confounding variables, *n*_*X*_ and *n*_*Y*_ are the corresponding sample sizes of exposure **X** and outcome **Y**, *J* is the number of genotyped SNPs. The error terms ***ε***_***X***_ and ***ε***_***Y***_ are independent noises from 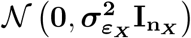 and 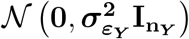 respectively. In model (3.1), *β*_0_ is the true causal effect and ***α*** represents the direct effect of the SNPs on the outcome not mediated by the exposure variable.

An external reference panel 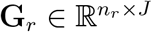 is chosen for estimating the LD matrix among SNPs, where *n*_*r*_ is the sample size of the chosen reference panel. The R package named *MR*.*LDP* is available to generate genotyped matrices **G**_***X***_, **G**_***Y***_ and **G**_***r***_. We fix *n*_*X*_ = *n*_*Y*_ = 20000 and *n*_*r*_ = 2500. The total number of SNPs is *J* = 300. For confounding variables, each column of **U**_***X***_ and **U**_***Y***_ is sampled from a standard normal distribution while each row of corresponding coefficients 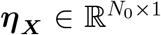 and 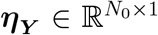 of confounding variables is obtained from a multivariate normal distribution 𝒩 (**0, *S***_***η***_) where the diagonal elements of ***S***_***η***_ ∈ ℝ^2×2^ are 1 and the off-diagonal elements are 0.8. We simulate the following three cases of idiosyncratic pleiotropy:

1. **case 1:** 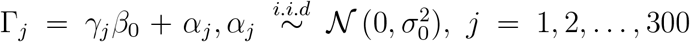. We randomly select {5%, 10%, 20%, 25%} of IVs so that their direct effects *α*_*j*_s have mean 0 and variance 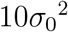.
2. **case 2:** 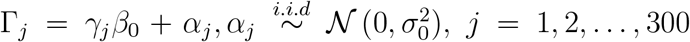. We randomly select {5%, 10%, 20%, 25%} of IVs so that their direct effect *α*_*j*_s have mean 10*σ*_0_ and variance 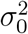.
3. **case 3:** 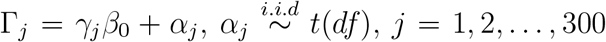, the values of freedom *df* are {10, 15, 20}.

The 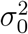 in case 1 and case 2 is controlled by the heritability *h*_***α***_ due to systematic pleiotropy, 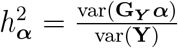 which is set to be 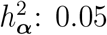 and 0.07 respectively. The signal magnitude for ***γ*** is chosen such that the heritability 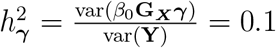 The true causal effect *β*_0_ is set to be 0.2.

We first run single-variant genetic association analysis for the exposure and the outcome respectively, and then we obtain the summary-level statistics 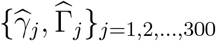 with their corresponding standard errors 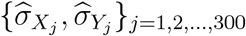 for all the three cases. Then we use the summary-level data to conduct MR analyses using the proposed RBMR, MR-LDP, MR-Egger, RAPS, GSMR and IVW methods. As the prerequisite for MR-Egger, RAPS and IVW methods is that the instrumental variables are independent of each other, we adopt a step-wise GSMR method to remove SNPs with LD structure. We repeat such experiment for 100 times.

The simulation results are shown in Figure 3.1. In all the three cases considered, we find that the proposed RBMR and MR-LDP methods are more stable than the other four methods: RAPS, GSMR MR-Egger and IVW. However, we found that our proposed RBMR method has smaller bias and root mean square error (RMSE) than the MR-LDP method. More detailed results are provided in the Supplementary Materials.

**Figure 3.1:**
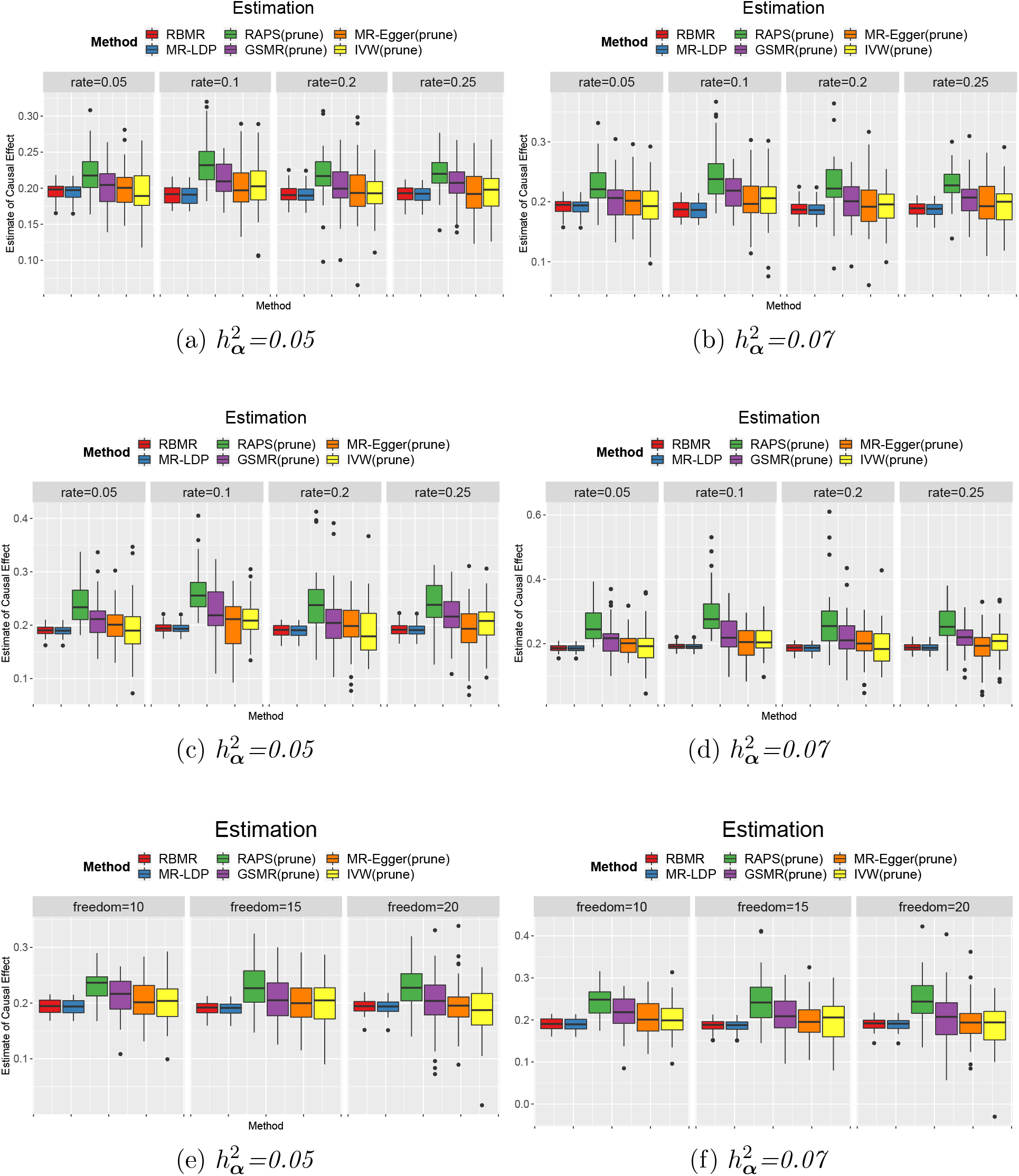
Comparisons of MR methods at heritability level of 0.05 and 0.07: The Figure (a), (c) and (e) represent comparisons of the causal estimates 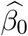 of RBMR, MR-LDP, RAPS, GSMR, MR-Egger and IVW methods at heritability level of 0.05 for three cases of idiosyncratic pleiotropy, respectively. The Figure (b), (d) and (f) represent comparisons of the causal estimates 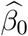 of RBMR, MR-LDP, RAPS, GSMR, MR-Egger and IVW methods at heritability level of 0.07 for three cases of idiosyncratic pleiotropy, respectively.

## 4. Real Data Analysis

In this section, we analyzed three real data sets to demonstrate the performance of our proposed method. The 1000 Genome Project Phase 1 (1KGP) is used as the reference panel to compute the LD matrix (Consortium et al., 2012). We first analyze two benchmark data sets commonly used for method comparison purpose, then we will estimate the causal effect of coronary artery disease (CAD) on the risk of critically ill COVID-19 outcome defined as those who end up on respiratory support or die from COVID-19.

The first benchmark data analysis is based on the summary-level data sets from two non-overlapping GWAS studies for the coronary artery disease (CAD), usually referred to as the CAD-CAD data. The true causal effect should be exactly one. The selection data set is from the Myocardial Infarction Genetics in the UK Biobank, the exposure data is from the Coronary Artery Disease (C4D) Genetics Consortium (Consortium et al., 2011), and the outcome data is from the transatlantic Coronary Artery Disease Genome Wide Replication and Meta-analysis (CARDIoGRAM) (Schunkert et al., 2011). We first filter the genetic variants using the selection data under different association *p*-value thresholds (*p*-value ≤ 1 × 10^*−*4^, 5 × 10^*−*4^, 1 × 10^*−*3^). Then we applied our proposed RBMR method and the MR-LDP to all the selected and possibly correlated SNPs by accounting for the LD structure explicitly. We applied the GSMR, IVW, MR-Egger and MR-RAPS methods using the independent SNPs after LD pruning because those methods require independent SNPs. We obtain causal effect point estimates and the corresponding 95% confidence intervals (CI) as shown in Figure 4.1(a). We found that our proposed RBMR method outperforms other methods because it has the smallest bias and shortest confidence intervals for a range of *p*-value thresholds.

**Figure 4.1:**
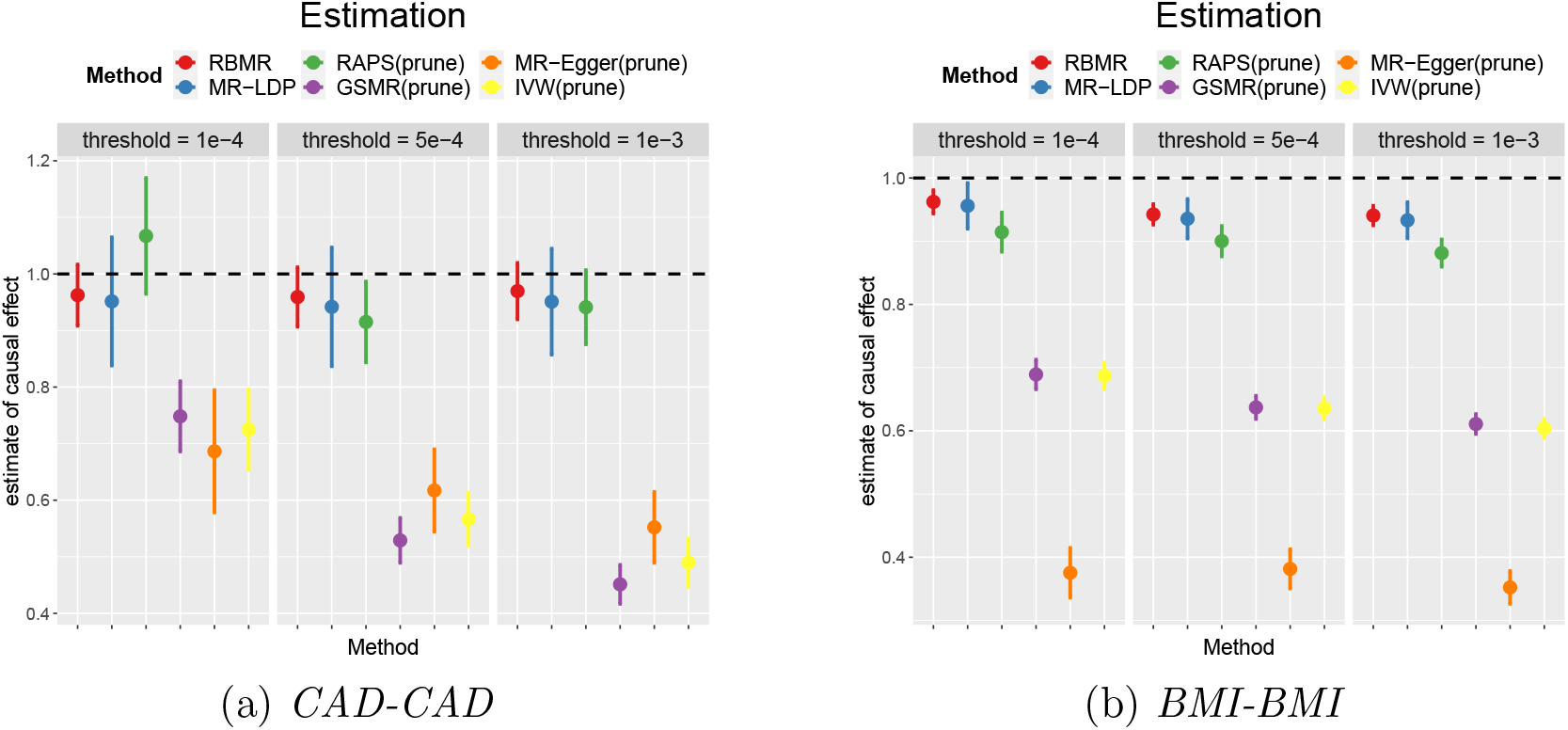
The results of CAD-CAD and BMI-BMI using 1KGP as the reference panel with shrink-age parameter λ = 0.15 and screening the corresponding SNPs under three thresholds (p-value ≤ 1 × 10^−4^, 5 × 10^−4^, 1 × 10^−3^),, RBMR, MR-LDP, RAPS, GSMR, MR-Egger and IVW methods use SNPs selected to calculate the casual effect estimate 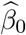.

To further investigate the performance of our proposed RBMR method, we consider the case that both the exposure and outcome are body mass index (BMI). We select SNPs based on previous research (Locke et al., 2015). The exposure is the BMI for physically active men and the outcome is the BMI for physically active women, both are of European ancestry (https://portals.broadinstitute.org/collaboration/giant/index.php/GIANT_consortium_data_files#2018_GIANT_and_UK_BioBank_Meta_Analysis_for_Public_Release). The point estimates and the corresponding 95% confidence intervals are shown in Figure 4.1(b). We found that our proposed RBMR method has smaller bias than other competing methods. More numerical results are provided in the Supplementary Materials.

We apply our proposed RBMR method together with other competing methods to estimate the causal effect of CAD on the risk of critically ill coronavirus disease 2019 (COVID-19) defined as those who end up on respiratory support or die from COVID-19. Specifically, the selection data set is the Myocardial Infraction Genetics in the UK Biobank and the exposure data set is from Consortium et al. (2011). The outcome is obtained from Freeze 5 (January 2021) of the COVID-19 Host Genetics Initiative (COVID-19 HGI) Genome-Wide Association Study (Initiative et al., 2020) (https://www.covid19hg.org/results/). The data combines the genetic data of 49562 patients and two million controls from 46 studies across 19 countries (Initiative et al., 2021). We mainly consider the GWAS data on the 6179 cases with critical illness due to COVID-19 and 1483780 controls from the general populations in our analysis. We use the selection data with *p*-value ≤ 1 × 10^*−*4^ threshold to select genetic variants as IVs. As shown in Figure 4.2(a), we found a significant effect of CAD on the risk of critically ill COVID-19 using our RBMR method (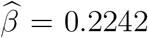, *p-*value = 0.0273, 95% CI = [0.0834, 0.3648]), MR-LDP (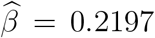, *p-*value = 0.0325, 95% CI = [0.0773, 0.3621]), RAPS (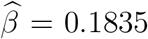, *p-*value = 0.0412, 95% CI = [0.0074, 0.3596])and MR-Egger (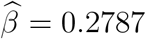, *p-*value = 0.012, 95% CI = [0.0607, 0.4967]) However, the results of GSMR (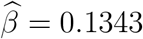, *p-*value = 0.0558, 95% CI = [− 0.0034, 0.2719]) and IVW (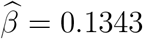, *p-*value = 0.0700 95% CI = [− 0.0111, 0.2796]) are not significant (*p*-value *>* 0.05). Although MR-LDP and RBMR give similar point estimate, however, our RBMR is more accurate as its confidence interval is slightly shorter and its *p*-value is more significant.

**Figure 4.2:**
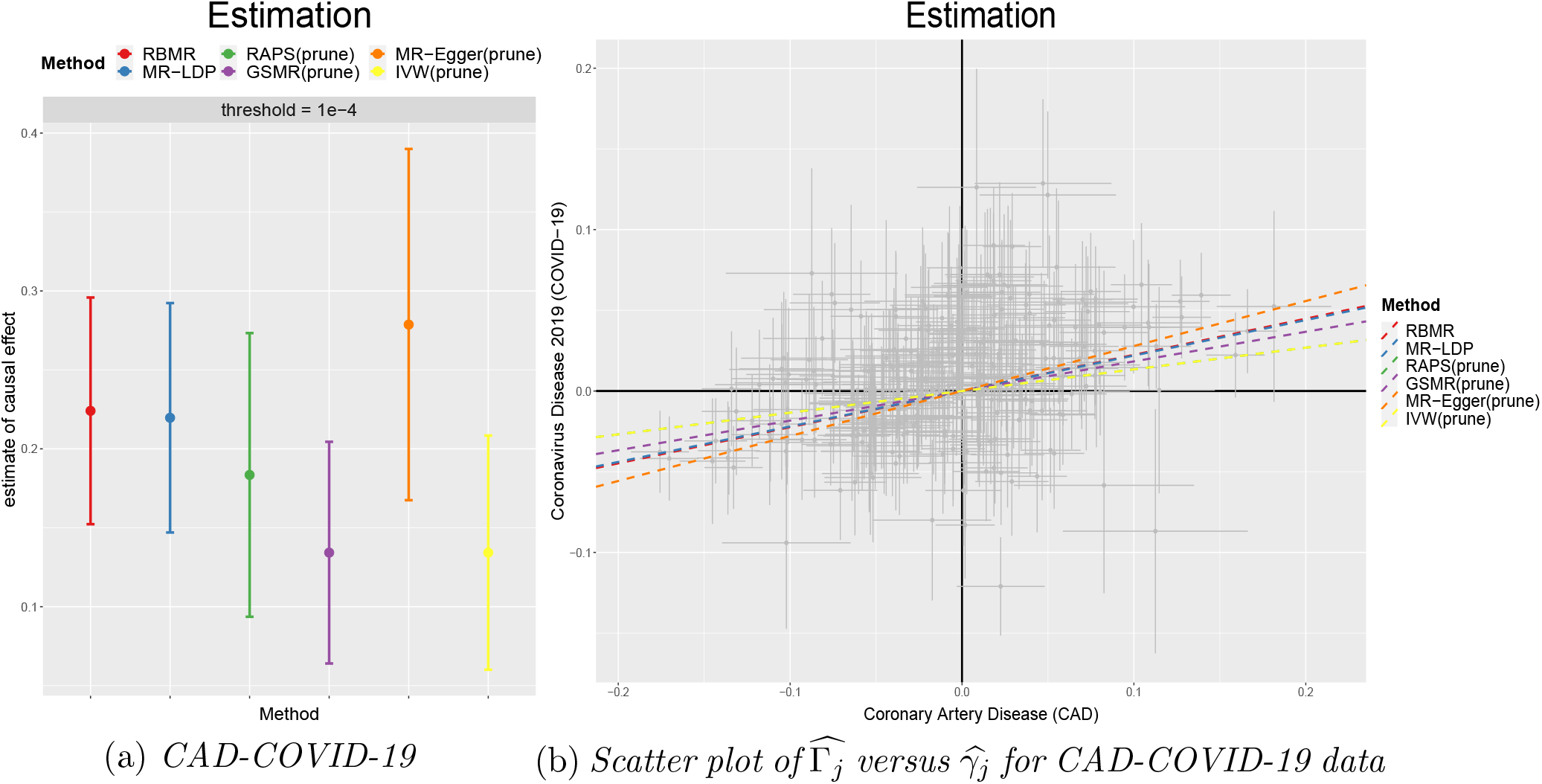
The results of CAD-COVID-19 using 1KGP as the reference panel with shrinkage parameter λ = 0.15 and screening the corresponding SNPs under thresholds (p-value ≤ 1 × 10^−4^), RBMR, MR-LDP, RAPS, GSMR, MR-Egger and IVW methods use SNPs selected to calculate the casual effect estimate 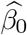. Each point of scatter plot in Figure (b) is augmented by the standard error of 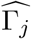 and 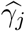 on the vertical and horizontal sides. Dashed lines are the slopes fitted by six methods.

## 5. Discussion

In this paper, we propose a novel two-sample robust MR method RBMR by accounting for the LD structure, systematic pleiotropy and idiosyncratic pleiotropy simultaneously in a unified framework. Specifically, we propose to use the more robust multivariate generalized *t*-distribution rather the less robust Gaussian distribution to model the direct effects of the IV on the outcome not mediated by the exposure. Moreover, the multivariate generalized *t*-distribution can be reformulated as Gaussian scaled mixtures to facilitate the estimation of the model parameters using the parameter expanded variational Bayesian expectation-maximum algorithm (PX-VBEM). Through extensive simulations and analysis of two real benchmark data sets, we found that our method outperforms the other competing methods. We also found that CAD is associated with increased risk of critically ill COVID-19 outcome using our RBMR method.

We make the following two major contributions. First, our method can account for the LD structure explicitly and thus can include more possibly correlated SNPs to reduce bias and increase estimation efficiency. Second, our RBMR method is more robust to the presence of idiosyncratic pleiotropy. One limitation of our proposed method is that it cannot handle correlated pleiotropy where the direct effect of the IV on the outcome might be correlated with the IV strength. We leave it as our future work.

## Data Availability

All data are publicly available as indicated or provided in the article.

## Acknowledgements

We would like to thank the GIANT Consortium and COVID-19 host genetics initiative for providing publicly available summary statistics to support our analysis.

